# Understanding motivations of older women to continue or discontinue breast cancer screening

**DOI:** 10.1101/2025.01.30.25321380

**Authors:** Susan C. Weller, Monique R. Pappadis, Shilpa Krishnan, Marsja R. Stearnes, Kristin M. Sheffield, Alai Tan, Jeffrey Z. Qi, James S. Goodwin

## Abstract

**Background:** Breast cancer screening guidelines indicate screening in women over 75 years of age is optional, depending upon patient health and preferences.

**Objectives:** To link experiences and perceptions of older women concerning screening to their intention to continue or discontinue screening.

**Design:** Qualitative comparative study, comparing continuers and discontinuers.

**Setting:** Community-residing adults.

**Participants:** A purposive sample (n=59) with equal representation of White, Black, and Hispanic women by age (70-74 years and 75 and older) and educational level (≤12 grade and >12 grade).

**Measurements:** In-depth qualitative interviews explored women’s perceptions of mammograms, the benefits and risks of screening, and personal screening experiences. Interviews were coded and quality-checked by two or more coders. A qualitative comparative analysis (QCA) was used to identify combinations of personal characteristics and themes linked to the intention to continue (n=32) or discontinue (n=27) screening.

**Results:** Personal experience themes were strongly linked to the intention to continue or discontinue. Women who mentioned recent screening (within three years) and either a spontaneously mentioned cancer story concerning a friend or family member or a doctor’s screening recommendation intended to continue screening (91% true positive rate, model sensitivity). Women who did not schedule screening and who did not mention a cancer story or a doctor’s recommendation (or neither) intended to discontinue screening (81% true negative rate, model specificity). These themes transcended differences in race/ethnicity, age, and educational level.

**Conclusion:** Continuation of breast cancer screening in older women is motivated by their personal screening history combined with cancer experiences and/or a doctor’s screening recommendation.

## INTRODUCTION

Breast cancer screening guidelines have converged on the age when women should start breast cancer screening, but ambiguity remains concerning when to stop. The US Preventive Services Task Force (USPSTF) recently broadened the age for screening, adding women 40-49 years of age, and now recommends screening for women 40-74 years of age, noting insufficient evidence of a benefit in women 75 and older.^1^ The American Cancer Society (ACS) and the American College of Obstetricians and Gynecologists (ACOG) recommend screening beginning at 40 and continuing while women are in good health with a life expectancy of 10 or more years.^2–4^ Screening of women who are older or have limited life expectancy can expose them to unnecessary treatment that may not prolong their life.^5–7^ By age 80, the average woman’s life expectancy is less than 10 years^8^ suggesting that discontinuation of screening would be appropriate. However, the receptivity of older women to discontinuation of screening is unclear, since half of women older than 75^9^ and many women with limited expectancy continue to be screened.^10,11^ To facilitate discussions with older women about screening, this paper explores women’s rationales for continuation and discontinuation of breast cancer screening.

Breast cancer incidence increases with older age and is highest in women 65 and older. Incidence peaks for women in their 70s, then decreases slightly, but remains high until their mid-80s.^12^ Mortality due to breast cancer increases steadily with older age,^13^ but can be reduced with mammography screening for breast cancers. Clinical trials demonstrate that screening reduces breast cancer mortality 14% in women 50-59 years, 33% in those 60-69 years, and 20% in women 70-74 years of age.^14^ Although incidence remains high for women aged 75 and older, no one in that age range was enrolled in the clinical trials – thus, there is no evidence for or against the effectiveness of mammography screening for women aged 75 and older.

Screening declines from almost three-quarters of women under 75 reporting screening to half of women over 75 reporting screening.^9^ For women in their 70s, shared decision-making between doctor and patient can help guide the decision to continue or discontinue screening.^1–3^ Informed and shared decision-making are patient-centered practices that inform patients about options and possible outcomes to help them make choices that best match their preferences.^15–17^ A patient-centered perspective is particularly important for breast cancer screening in older women because mammography may have a net benefit for some patients but not others and because older women may assign different values to the benefits and risks of screening.^18,19^ Studies suggest that women care more about early detection of cancer than about the negative aspects of screening, such as detection of cancers that would not affect mortality.^20,21^ Although some older women may insist on continuing breast cancer screening,^22–24^ screening decisions may be influenced by a doctor’s recommendation, personal screening history, and the reassurance that screening may offer.^22,25–27^

This study explores factors associated with the decision to continue or discontinue screening in women in the age range where discontinuation tends to occur (70 and older). However, this study departs from previous descriptive studies of positive and negative aspects of mammogram screening and screening preferences by instead identifying combinations of key factors that predict the intention to continue or discontinue screening. A qualitative comparative analysis (QCA)^28,29^ of in-depth interviews identified themes and combinations of themes uniquely linked to continuation or discontinuation of breast cancer screening. QCA is *similar to* logistic regression, bridging qualitative and quantitative analyses, but is based on set and subset relations rather than quantitative measures or probability.^30^ A strength of QCA is its ability to identify complex interactions with small sample sizes, allowing for asymmetry in prediction, in case the factors predicting continuation differ from those predicting discontinuation. Specifically, this study explored older women’s perceptions of breast cancer screening, to discover whether the decision to continue screening is driven primarily by a physician’s recommendation and if not, what other factors contribute to a woman’s screening intention.

## METHODS

This study was part of a larger project examining older women’s motivations for screening, their willingness to discontinue screening,^24^ their understanding of overdiagnosis,^31^ and their preferences for communication about screening,^32^ but departs from prior studies in attempting to identify the pathway between experiences and the intention to screen.

### Participants

Women 70 years and older with no prior history of breast cancer were recruited from a local geriatric clinic, senior community living facilities, senior public housing, local community centers, and churches in southeast Texas. Because of disparities in screening,^33–35^ a purposive sample was sought with equal representation across age, race/ethnicity, and educational level.^36^ Approximately five women were selected from each of the 12 categories formed by younger-old/older-old ages (70-74 vs. 75+), lower/higher education (≤12 vs. >12 grade), and race/ethnicity (White, Black, Hispanic). The sample size was adequate as a sample size of 5 can capture ideas with a 30% or higher population prevalence and a sample of 16 (combining subgroups) can capture ideas with a 10% or higher population prevalence (with 80% confidence);^37^ a comparison of ideas and themes from continuers and discontinuers aggregated across demographic subgroups would ensure capturing the most salient ideas and themes.^38^ The study was approved by the University’s Institutional Review Board; participants provided oral informed consent (audiotaped) and received a $25 gift card for their time. Women were recruited between 8/27/13 and 3/26/18. This study integrates their data with a new analysis.

### Procedure

Women were asked a series of open-ended questions about mammogram screening (See Supplement, Table A). A first set of questions covered general information about mammograms: their description, purpose, benefits, and risks. A second set of questions explored the decision process. All interviews were audiotaped, transcribed, and coded.

### Analysis

Themes were identified by repetitions within and across transcripts.^39^ For each substantive theme, the interviewer (MS) did the first coding and drafted a codebook in NVivo 10.^40^ Then, a second and third person (MRP, SK) independently coded themes, and team members (MS, AT, SW, MRP, SK) reviewed transcripts for codes relevant to each theme and iteratively revised the codebook. Codes were quality-checked via content analysis of transcripts using specific keywords and phrases (“keywords in context”)^41^ with review and validation by team members. Content for the decision process (who initiated screening, did the doctor recommend screening, mention of a personal cancer story, mention of screening reminders, and whether screening will be continued) was described by codes and quotes.

To see if narratives of women intending to continue screening differed from those who did not, the salience of each theme/code was estimated within each group and a qualitative comparative analysis (QCA) was conducted to identify key factors in the decision pathway to continuation/discontinuation. Responses of each woman were represented in a spreadsheet as a dichotomous profile representing the presence/absence of themes mentioned when talking about mammograms. Theme salience for discontinuers/continuers was estimated with the proportion in each group mentioning each theme/code^38,41,42^ and compared with a chi square test.^43^

QCA identified key factors (“prime implicants”)^41^ linked to the intention to screen by comparing the profiles of thematic code usage of individuals intending to continue or discontinue. QCA^28–30,41^ considers all possible combinations of codes and logical relationships are used to link thematic codes to group membership (implemented in Tosmana^44^). QCA uses formal Boolean logic (set and subset relations) to identify important distinguishing factors between subgroups. For example, if (1) all women (young and old) with a doctor’s recommendation wanted to continue screening and (2) all women (young and old) without a doctor’s recommendation wanted to discontinue screening, then a doctor’s recommendation distinguishes continuers from discontinuers, while age does not.

## RESULTS

### Sample Characteristics

Fifty-nine English-speaking women were interviewed for this study: 28 were 70-74 years old and 31 were 75 and older; 24 were White, 21 Black, and 14 Hispanic; and 29 had lower and 30 had higher educational levels. Although the sampling target was for five women in each subcategory, some categories (e.g., older Hispanics with higher educational levels) were difficult to locate and some were slightly over-represented (e.g., White women with higher educational levels). Most women were recruited from community settings and very few rated their health poorly. The interviews lasted between 45 to 60 minutes.

### Intention to Continue or Discontinue Screening

Approximately half of the women (n=32) expressed an intention to continue screening and half (n=27) did not. Those who were unsure about continuing (n=1) and those who stated their intention to follow their doctor’s recommendation to discontinue screening (n=4) were coded as discontinuers. Those who reported they would follow their doctor’s recommendation to continue screening were considered continuers (n=2). The decision to continue/discontinue screening was associated with older age (67% of discontinuers vs. 41% of continuers, p<.05), but not education or race/ethnicity (Table 1).

**TABLE 1:** DEMOGRAPHIC CHARACTERISTICS BY SCREENING INTENT.

|  | DISCONTINUE<br>(n=27) | CONTINUE<br>(n=32) |
| --- | --- | --- |
| **Demographics** |  |  |
| Age (75+ yrs)* | 67% (18/27) | 41% (13/32) |
| Education (> High School) | 48% (13/27) | 53% (17/32) |
| Race/Ethnicity |  |  |
| White | 44% (12/27) | 38% (12/32) |
| African-American | 33% (9/27) | 38% (12/32) |
| Hispanic | 22% (6/27) | 25% (8/32) |
| Community Recruitment | 86% (19/22) | 90% (26/29) |
| Self-Rated Health (very poor) | 7% (2/27) | 0% (0/32) |

\* p< .05

Many women said they would continue screening as long as they lived (“*As long as I live*,” “*I guess until I die,*” or “*As long as I am able*”) or that they did not intend to stop (“*I would not consider stopping*” or “*I don’t intend to stop*”). Some linked continuation to a doctor’s recommendation, for example: “*Probably indefinitely unless the doctor would cut me off and say that it’s not necessary any longer.*”

Discontinuation was expressed simply as “*I figure I’m through having them*,” often linked to advanced age: “*I’m 83 years old and I don’t feel like I really need one,*” or “*I probably won’t have one next year, just because I don’t think it’s necessary for somebody 76 years old next year, and I’ve never had any problems at all,*” or to a doctor’s recommendation in the context of age, “*The suggestion was when I first started out to get them annually, and then later. I don’t remember exactly when we moved it forward, and then after 60, probably five years. Then I have nothing after maybe 70, and I haven’t had any.*”

### General Descriptions, Purpose, Benefits, and Risks of Mammograms

Most of the women were very familiar with mammograms. Only two women had never had one and one additional woman said that she had one, but described a screening procedure that may have been an MRI. Narratives concerning mammogram descriptions, purpose, benefits, and risks were fairly consistent across age, educational, and racial/ethnic groups with few meaningful differences between discontinuers and continuers (Table 2). Women described mammograms as an X-ray or machine; an image, picture, or photo of the breast; or more generally as a test, exam, check, or screen of the breast. The purpose was to detect cancer; detect lumps or nodes; or detect abnormalities. Benefits were to detect cancer (themes describing the purpose of screening often were repeated and emphasized), to detect lumps/nodes/abnormalities; to detect problems early to get timely treatment; to live longer; and to gain health reassurance. Some reported that mammograms had no risk, while others reported risk due to radiation exposure. Pain/discomfort was reported as a risk, as was the possibility of an inaccurate reading. Only two of the 15 codes appeared to differ in salience between discontinuers and continuers. Discontinuers were somewhat more likely to describe mammograms as a “breast X-ray” (56% vs. 35%, p<.18) and cite the benefit of “cancer detection” (48% vs. 28%, p<.20).

**TABLE 2:** MAMMOGRAM CHARACTERISTICS AND THEMATIC CODES BY INTENT TO CONTINUE SCREENING.

| Theme, code (*examples*) | DISCONTINUE | CONTINUE |
| --- | --- | --- |
| **Description** |  |  |
| Breast X-ray (*"It's an X-ray of your breasts."*) | 56% (15/27) | 36% (11/31) |
| Breast exam (*"test of the breast"*) | 26% (7/27) | 23% (7/31) |
| Breast image (*"when they do the image of the breast"*) | 22% (6/27) | 32% (10/31) |
| **Purpose** |  |  |
| Detect cancer (*"To find out whether you have cancer."*) | 62% (16/26) | 66% (21/32) |
| Detect lump (*"To see if you have any growths or detect lumps that shouldn't be there."*) | 42% (11/26) | 28% (9/32) |
| Detect abnormality (*"To detect tumors or abnormalities in the tissue."*) | 19% (5/26) | 19% (6/32) |
| **Benefits** |  |  |
| Detect cancer (*"Test of the breast to make sure there's no cancer cells or cancer."*) | 48% (13/27) | 28% (9/32) |
| Detect lump/abnormality (*"To detect any abnormalities in your breast."*) | 26% (7/27) | 31% (10/32) |
| Timely treatment (*"to catch it early"* or *"catch it in time"*) | 26% (7/27) | 38% (12/32) |
| Health Reassurance (*"To tell if you're alright or if you're not"*) | 22% (6/27) | 25% (8/32) |
| Live longer (*"keep myself healthy and alive for my children"*) | 7% (2/27) | 9% (3/32) |
| **Risks** |  |  |
| No risk (*"I never heard of any risks"*) | 26% (7/27) | 38% (12/32) |
| Radiation/cancerous (*"The possibility of getting cancer from the X-ray."*) | 30% (8/27) | 22% (7/32) |
| Pain or discomfort (*"women don't like to be squeezed. They always said it hurts so hard."*) | 22% (6/27) | 19% (6/32) |
| Inaccurate reading (*"False readings"* or *"mistakes"*) | 15% (4/27) | 6% (2/32) |

### Decision Process for Screening

Discussions about the screening process revealed personal experiences that were more strongly linked to the intention to continue/discontinue screening than were general descriptions of mammograms, including the purpose, benefits, and risks of mammograms. Self-initiation of screening appointments, having a doctor’s recommendation, recent screening (≤3 years), and spontaneous mention of a cancer story involving a friend or family member showed large, statistically significant differences in salience between the two groups (Table 3).

**TABLE 3:** DECISION PROCESS THEMATIC CODES BY INTENT TO CONTINUE SCREENING.

| Theme, code (*examples*) | DISCONTINUE | CONTINUE |
| --- | --- | --- |
| **Decision Process** |  |  |
| Doctor initiated screening<br>(*"It's usually set-up with my primary care that she'll tell me, 'this year you'll go for eye, or bone density, or mammogram,' you know, and I just know that it's...I go every spring, and that's it."*) | 74% (20/27) | 69% (22/32) |
| Self-Initiated screening*<br>(*"Sometimes I remind my doctor that it's time."*) | 7% (2/27) | 31% (10/32) |
| Reminder<br>(*"I just got the letter last month I think it was."*) | 19% (5/27) | 22% (7/32) |
| Personal cancer story**<br>(*"There is breast cancer in my family,"* or *"...on my father's side of the family there's been all kinds of cancers."*) | 33% (9/27) | 75% (24/32) |
| Doctor recommended screening (verbally) **<br>(*"My doctor is the one that says,"* or, *"I just went along with it."*) | 19% (5/27) | 69% (22/32) |
| Recent screening, screening within past three years** | 41% (11/27) | 91% (29/32) |

\* p< .05, \*\* p< .001

Women described the initiation of screening as doctor-initiated, self-initiated, or both. Those who intended to continue screening were more likely to report ***self-initiation of screening appointments*** (31% vs. 7%, p<.05). Although it was expected that screening might be influenced by a doctor scheduling an appointment or by a call/letter reminder, only a ***doctor’s verbal recommendation*** influenced intent. A doctor’s recommendation was reported much more often by those who intended to continue screening (69% vs. 19%, p<.001). Those who desired to continue screening were more likely to have ***had a recent mammogram*** (91% vs. 41%, p<.001) and were more likely to ***spontaneously mention a cancer story involving family or friends*** (75% vs. 33%, p<.001).

### Themes Linked to the Decision to Continue

Of the 21 thematic codes (Tables 2 and 3) and three demographic characteristics (age, race/ethnicity, educational level), only five factors were associated with the intent to continue screening: younger age, reporting a cancer story involving family or friends, self-initiation of screening, recent screening, and receiving a doctor’s recommendation (Supplement, Table B). Of the 32 possible combinations of the presence/absence of the five factors, only 19 occurred in the 59 cases and only four of the themes (excluding age) were linked to the intention to continue/discontinue. Table 4 shows the 19 combinations of the factors that occurred in the 59 narratives, the case counts for each distinct profile of themes, and the classification rules linked to continuation/discontinuation. For example, there were 15 women who reported a personal cancer story (+), a doctor’s recommendation (+), and recent screening (+); 14 (6+7+1) wanted to continue and 1 to discontinue.

**TABLE 4:** PROFILES FOR ALL 59 CASES & PREDICTION RULES.

| Profile | Older Age | Initiated Screen | Cancer Story | Dr. Recom | Recent Screen | Continue # Cases (n=32) | Discontinue # Cases (n=27) |
| --- | --- | --- | --- | --- | --- | --- | --- |
| **CONTINUE RULE#1:**<br>**Recent Screen + Cancer Story + Doctor's Recommendation** |  |  |  |  |  |  |  |
|  | + | - | + | + | + | 6 | 1 |
|  | - | - | + | + | + | 7 | 0 |
|  | - | + | + | + | + | 1 | 0 |
| **CONTINUE RULE#2:**<br>**Recent Screen + Cancer Story - No Doctor's Recommendation** |  |  |  |  |  |  |  |
|  | + | + | + | - | + | 5 | 0 |
|  | - | + | + | - | + | 2 | 0 |
|  | + | - | + | - | + | 1 | 1 |
| **CONTINUE#3:**<br>**Recent Screen + Doctor's Recommendation - No Cancer Story** |  |  |  |  |  |  |  |
|  | - | + | - | + | + | 1 | 0 |
|  | + | + | - | + | + | 0 | 1 |
|  | - | - | - | + | + | 6 | 0 |
|  | + | - | - | + | + | 0 | 2 |
| **DISCONTINUE RULE#1:**<br>**No Initiated Screening - No Cancer Story - No Doctor's Recommendation** |  |  |  |  |  |  |  |
|  | + | - | - | - | - | 0 | 6 |
|  | + | - | - | - | + | 0 | 3 |
|  | - | - | - | - | - | 0 | 2 |
|  | - | - | - | - | + | 0 | 3 |
| **DISCONTINUE RULE#2:**<br>**No Initiated Screening - No Cancer Story + Doctor's Recommendation** |  |  |  |  |  |  |  |
|  | + | - | - | + | - | 1 | 0 |
|  | - | - | - | + | - | 0 | 1 |
| **DISCONTINUE RULE#3:**<br>**No Initiated Screening + Cancer Story - No Doctor's Recommendation** |  |  |  |  |  |  |  |
|  | + | - | + | - | - | 0 | 4 |
|  | - | - | + | - | - | 1 | 2 |
| **Unaccounted for** |  |  |  |  |  |  |  |
|  | - | + | + | - | - | 1 | 1 |
| **TOTAL** |  |  |  |  |  | 32 | 27 |

The intention to continue screening was motivated by recent screening, a cancer experience, and a doctor’s recommendation, regardless of age. Women with all *three* of these factors intended to continue (Rule #1). All continuers had a recent mammogram screening, but it was not sufficient to predict continued screening. The power of a personal cancer experience (Rule #2) *or* a doctor’s recommendation to continue screening (Rule #3) was clear as women of all demographic groups were motivated to continue screening if they had recent screening *and* either a cancer story or a doctor’s recommendation or both. These three factors correctly classified 29/32 cases desiring to continue (91% true positives).

Similarly, the intention to discontinue was characterized by the absence of a cancer story, absence of a doctor’s recommendation, and the absence of self-initiated screening (Rule #4). All who intended to discontinue had not self-initiated screening in the past, although they may or may not have been recently screened. Failure to self-initiate screening combined with the absence of a doctor’s recommendation (Rule #5) and/or a personal cancer story/experience (Rule #6) predicted intent to discontinue. A doctor’s recommendation to screen was ignored by women who did not self-initiate screening and had no motivating personal cancer story. These factors correctly classified 22/27 cases desiring to discontinue (81% true negatives). One profile (two women who self-initiated screening, reported a cancer story, but did not have a doctor’s recommendation; bottom, Table 4) remained unaccounted for (one woman wanted to continue and one wanted to discontinue). Since a classification rule accommodating this profile would count as either a correct continuation and an error for discontinuation or vice versa, we simply counted it as an error against both continuation and discontinuation predictions.

## DISCUSSION

Few studies describe the breast cancer screening preferences of older women, and those that do rarely link specific attributes and themes to actual screening or the intention to screen.^27,45–49^ This is the first study to directly link themes to the intention to screen. Themes identified in this study are similar to those identified in other studies,^22,25,46^ but in addition to listing salient themes, here we identified combinations of themes explicitly motivating the intention to continue or discontinue screening in older women. Continued screening is primarily a function of past screening behavior (recent screening), but past screening must be combined with a personal cancer experience and/or a doctor’s verbal recommendation to motivate future screening. Women who have been recently screened and who have had a friend or family member with cancer are likely to continue to screen with or without a doctor’s recommendation. For women who have been screened but who do not know someone with cancer, a doctor’s recommendation is the tipping point for continuing screening. While other studies on women in their 70s identified the importance of a doctor’s recommendation,^26^ this study shows a doctor’s recommendation to get screened only activates future screening when combined with recent screening.

Discontinuation of screening is similarly a function of past screening, but discontinuation is determined by whether a woman has self-initiated screening rather than whether she was recently screened. Women who did not self-initiate their screening appointments and who have not known someone with cancer and/or do not have a doctor’s recommendation are likely to discontinue screening. If a doctor does not give a recommendation for screening and the doctor’s office or health care system does not schedule screening, screening is likely to be discontinued. A woman can override this passive discontinuation option, however, by scheduling her own appointment, especially if she has known someone with cancer. Often, discontinuation of breast cancer screening seems to follow a passive path.

The role of older age in the decision to discontinue has been unclear. Some assume that older age may be a consideration for patients, as mammogram screening declines with age (with large decreases occurring at 75 and 80 years).^9,50^ However, older age has not been directly linked to a desire to discontinue.^25^ Older-old women in this study were more likely to express a desire to discontinue, but results indicated that other factors played more important roles in a woman’s decision to continue or discontinue screening than simply older age.

Women in this study were generally healthy, as few rated their health as poor. Poor self-rated health is linked to higher mortality^51,52^ and the two women who rated their health as poor intended to discontinue screening. There were no systematic racial/ethnic or educational differences in narratives linked to the intention to continue/discontinue breast cancer screening. These findings parallel those from a national survey that did not find race, age, or health status differences in utilization.^49^

Women in this study also were fairly well-informed about breast cancer screening. All women were able to describe a mammogram and its purpose. Previous studies described harms and risks of screening mammography, including possible unnecessary diagnostic procedures, costs, complications, as well as pain and anxiety.^1,18^ In this study, women reported pain, possible radiation risk, and the possibility of inaccurate results, but not risk from unnecessary treatment. Furthermore, these perceived risks did not motivate their decision to continue or discontinue. Risks of mammogram screening are sometimes cited as reasons to discontinue screening, but women appear to prioritize detection of cancer over potential risks.^18,25,53^ Thus, screening may offer the benefit of health reassurance (“better to know” or “peace of mind”).^20,25,53^ However, in this study the effects of perceived risks and benefits were surpassed by a personal cancer story, a story that may express a perceived higher personal risk for cancer. Experiences of friends and family may lead women to overestimate breast cancer incidence, mortality, and the predictive value of mammography – and thus, overestimate their own personal risk of being diagnosed with and dying from breast cancer.^18,54^

Physicians are an important source of information on screening and affect patient uptake of screening.^22,26,49^ In older women who have been recently screened, a provider can influence the decision to screen.^25,47^ This study supports the idea that a tipping point in the decision for many older women may be the recommendation – or lack of a recommendation – to get screened. Cessation may be a harder decision than continuing and discussion of cessation may even compromise patient-provider trust,^23^ but a simple reduction in reminders may reduce overuse in older patients.^25^

A limitation of this study is the possible inaccuracy in recollections of past behaviors and reports on future intentions. Women may decide what they want to do and then report information that supports that action. For example, a woman who wants to be screened may be more likely to remember and report that her doctor recommended it. Also, women were not followed prospectively to verify their intentions, although intention is a strong predictor of behavior and can serve as a proxy for actual behavior.^47^ Another limitation is that the study was based on a small regional sample and, as for most qualitative studies, results are suggestive and not definitive. In-depth exploration allows for the identification of new ideas and processes that need to be validated with an independent, representative sample. However, the replication of many themes in this study with those in prior studies^22,25,27,46,47^ adds evidence toward the validity of our findings.

A strength of the study was that QCA facilitated the understanding of complex relationships among factors linked to the intention to screen, winnowing down almost two dozen themes to a less than a handful. QCA bridges qualitative and quantitative approaches by combining qualitative data with a systematic analysis of cases.^55^ However, results can be sensitive to the selection of explanatory variables or the addition of more cases. Our analysis focused on the five factors that maximally discriminated continuers and discontinuers and, although QCA estimates a causal model, our models did not incorporate temporal relationships, as the outcome variable was intention to screen and not prospectively verified behavior. Never-the-less, QCA suggests the path between themes, experiences, and perceptions to the intention to continue or discontinue screening.

QCA offers a way to interpret patterns in qualitative data that is systematic and transparent. Our analysis systematically compared narratives of women expressing a desire to continue with those who might discontinue and estimated true positive and true negative classification rates for each group. QCA results, however, may not be unique: there may be more than one set of rules to account for response patterns. Classification of cases can be complicated and the trade-off is a more complex set of rules with greater accuracy versus a more parsimonious solution with reduced accuracy. Here, a few cases were counted as errors, when they could have been accommodated with extra classification rules. Thus, we opted for fewer rules that slightly reduced the true negative rate.

## Conclusion

Among the many positive and negative aspects of screening, only a few tend to motivate older women to continue or discontinue breast cancer screening. Foremost is their past screening behavior. Then, intentions are motivated by a personal cancer experience in a friend or relative and/or a doctor’s recommendation. Some may be ready to discontinue, especially those who delay or skip reminders, and may be most influenced by a discussion with their physician about continuing or discontinuing screening.

## Data Availability

All relevant data are within the manuscript and its Supporting Information files.

